# Sleep and seizure risk in epilepsy: Bed and wake times are more important than sleep duration

**DOI:** 10.1101/2022.08.05.22278453

**Authors:** Rachel E. Stirling, Cindy M. Hidajat, David B. Grayden, Wendyl J D’Souza, Katrina L. Dell, Ray Boston, Logan D. Schneider, Ewan Nurse, Dean Freestone, Mark J. Cook, Philippa J. Karoly

## Abstract

Sleep duration, sleep deprivation and the sleep-wake cycle are thought to play an important role in the generation of epileptic activity and may also influence seizure risk. Hence, people diagnosed with epilepsy are commonly asked to maintain consistent sleep routines. However, emerging evidence paints a more nuanced picture of the relationship between seizures and sleep, with bidirectional effects between changes in sleep and seizure risk in addition to modulation by sleep stages and transitions between stages. We conducted a longitudinal study investigating sleep parameters and self-reported seizure occurrence in an ambulatory at-home setting using mobile and wearable monitoring.

Forty-four subjects wore a Fitbit smartwatch for at least 28 days while reporting their seizure activity in a mobile app. Multiple sleep features were investigated, including duration, oversleep and undersleep, and sleep onset and offset times. Sleep features in participants with epilepsy were compared to a large (n=37921) representative population of Fitbit users, each with 28 days of data. For participants with at least 10 seizure days (n=29), sleep features were analysed for significant changes prior to seizure days.

A total of 3894 reported seizures (M = 88, SD = 130) and 17078 recorded sleep nights (M = 388, SD = 351) were included in the study. Participants with epilepsy slept an average of 2 hours longer than the average sleep duration within the general population. Just 1 of 29 participants showed a significant difference in sleep duration the night before seizure days compared to seizure-free days. However, 11 of 29 subjects showed significant differences between either their sleep onset (bed) or offset (wake) times prior to seizure occurrence. In contrast to previous studies, the current study found oversleeping was associated with a 20% increased seizure risk in the following 48h (p < 0.01), likely due to nocturnal seizures driving increased sleep durations.

Nocturnal seizures were associated with both significantly longer sleep durations and increased risk of a seizure occurring in the following 48h. Oversleeping only significantly contributed to increased seizure risk when participants were already in a high-risk (rather than baseline- or low-risk) state, according to their endogenous cycles of seizure likelihood.

Overall, the presented results demonstrated that day-to-day changes in sleep-duration had a minimal effect on reported seizures, while bed- and wake-times were more important for identifying seizure risk the following day. Oversleeping was linked to seizure occurrence, most likely due to nocturnal seizures driving oversleep. Wearables can be utilised to identify these sleep-seizure relationships and guide clinical recommendations or improve seizure forecasting algorithms.

## Introduction

The complex bi-directional relationship between epileptic seizures and sleep has been of interest to researchers for centuries.^1–3^ In 1885, Gowers observed that almost two-thirds of people with epilepsy experienced seizures exclusively during either the night or day.^2^ Since then, many studies have confirmed the existence of ‘pure sleep epilepsy’, albeit at a lower prevalence, and have highlighted that sleep, predominantly the stage of sleep, plays a role in epileptic activity.^1^ In seizure forecasting, sleep staging and duration has shown individual-specific predictive utility using long-term electrographic seizure records^4^ and wearables.^5^ However, the role of sleep in epilepsy remains unclear, as the impact of sleep often depends on the type of syndrome,^1,6,7^ the cerebral lobe of seizure origin^8^ and the type of treatment being used.^9^ Further elucidating this relationship via practical individual-specific avenues may aid in improving seizure management, clinical outcomes, and seizure forecasting.

Unforced sleep deprivation (undersleep) has traditionally been one of the most commonly reported factors associated with seizure occurrence, so much so that forced sleep deprivation has historically been used prior to EEG assessment to improve the diagnostic yield of seizures or interictal epileptiform discharges (IEDs).^10^ However, emerging evidence paints a more nuanced picture of the relationship between seizures and sleep duration, with too much (oversleep) and too little sleep (deprivation) both linked to seizure occurrence.^4,11–13^ Rajna and Veres^13^ showed in a long-term diary study that sleep deprivation (1.5 hours less than mean sleep duration) increased seizure risk by six-fold the following day compared to ‘normal’ sleep. Oversleeping (1.5 hours greater than mean sleep duration) also increased the likelihood of seizures by three-fold. Contrary to this, in 2021, Dell et al.^11^ showed in a long-term intracranial electroencephalogram (EEG) study that sleep deprivation (sleeping less than the 25th percentile or 1.13 hours below median) had no significant effect on the risk of a seizure (identified by intracranial EEG) in the following 48h, whereas oversleep (sleeping more than the 75th percentile, or 1.66 hours above median) reduced the risk of a seizure by 27% in the following 48 hours, for 10 people with refractory focal epilepsy. Forced sleep deprivation in shorter inpatient studies, on the other hand, has been shown to improve diagnostic yield of EEGs in some studies,^14^ but not others.^15^ In any case, forced sleep deprivation before EEG monitoring – often also accompanied with medication withdrawal – does not reliably reflect fluctuations in individuals’ sleep in their normal home environment. Nonetheless, there is clearly a relationship between sleep duration and seizures. However, the evidence is not clear as to whether increased or decreased sleep drives seizure risk. Some contradictory findings are likely due to the diverse duration and setting (i.e. inpatient vs at home) of sleep studies in epilepsy.

Circadian rhythms and the sleep cycle are also implicated in the occurrence of seizures and IEDs.^16,17^ In a small but seminal study, Pavlova et al.^18^ found that IEDs were related to both circadian rhythms and sleep stage in generalised epilepsy, with IEDs 14 times more likely to occur during NREM sleep than REM sleep or wakefulness, independent from environmental cues. Recently, these findings have been enhanced by further work investigating the relationship between NREM sleep and epileptic seizures,^7,16^ supporting the theory that epileptic discharges are related to the sleep/wake cycle rather than just circadian or behavioural patterns. Transitions from sleep-to-wake and wake-to-sleep are also important, as sleep onset and offset can precipitate seizures in some people with focal epilepsy.^11^ However, the timing of seizures in the sleep-wake cycle often depends on epilepsy syndrome and lobe of origin.^6,19,20^

These findings collectively demonstrate that existing research does not paint a clear picture of the relationship between epilepsy and sleep. Clinicians often overcome this by counselling their patients on the general importance of sufficient sleep.^21^ However, the lack of objective data means that this advice may be unhelpful for some people. The current work investigated relationships between sleep and self-reported seizures using a wearable smartwatch and mobile seizure diaries, as non-invasive devices enable long-term recordings in an at-home setting with minimal disruption of daily life. Here we aim to untangle decade-long beliefs in the sleep and epilepsy field and provide clearer guidelines for sleep counselling in epilepsy management.

## Materials and Methods

We recruited 44 adults (31 female) from a commercial database (Seer Medical, Australia) who wore a smartwatch for at least 28 nights and reported at least one seizure in the Seer app (Seer Medical, Australia). Participants were 20 - 71 years of age, all with a diagnosis of epilepsy. Participants who reported their epilepsy syndromes had a mix of Focal, Multi-focal, Genetic Generalised Epilepsy or Generalised diagnoses (Supplementary Table 1). All participants provided informed consent for the study (St Vincent’s Hospital HREC 009/19).

## Data collection

Continuous data were collected via mobile and wearable devices for at least 28 days and up to 5 years. All participants wore a smartwatch (Fitbit, Alphabet Inc., USA) and manually reported their seizure times in a mobile diary app (Seer App). Sleep data (sleep stages and durations) was provided by Fitbit using a proprietary sleep algorithm, which estimates sleep stages (categorised into Light, Deep and REM sleep) using a combination of movement (via accelerometery) and heart rate (via photoplethysmography) patterns, including heart rate variability. According to recent studies, Fitbit adequately estimates total sleep duration, sleep onset and offset, and durations of sleep stages in most cases^22,23^; however it is not yet a suitable substitute for gold-standard polysomnography, thus sleep stage analyses have primarily been included as supplemental data in this work. Using Fitbit, light sleep approximately corresponds to N1/N2 stages, deep sleep approximately corresponds to N3 stages, and REM sleep marked by Fitbit reflects the more autonomically unstable stage of sleep (still referred to as ‘REM’ in this study for clarity).

## Pre-processing data

To evaluate the relationship between sleep and seizures, features were derived from unprocessed sleep stage data from the Fitbit API. The sleep stages and durations were transformed into six daily features similar to previous work,^11^ which allowed us to compare results:

1. Total sleep duration (longest overnight sleep, excluding naps)
2. Sleep onset time (bedtime)
3. Sleep offset time (waketime)
4. Total duration spent in Deep / Light / REM stages during sleep
5. Proportion of duration spent in Deep / Light / REM stages during sleep
6. Total seizures (sum of seizures from sleep onset to sleep onset the following day)

## Statistics

This study assessed four main relationships: (1) Sleep variables (total sleep duration and duration/proportion of Deep / Light / REM / Wake states overnight) in people with epilepsy compared to the general population, (2) Sleep duration and duration trend prior to seizure days compared to seizure-free days; (3) Oversleep and undersleep (also referred to as sleep deprivation in the literature) leading up to seizure days compared to seizure-free days; and (4) Bedtime and waketime on seizure days compared to seizure-free days. The statistical tests used in each of these results sections are described in the sections below.

### Sleep in epilepsy compared to the general population

To evaluate each of the sleep variables of interest for the epilepsy cohort compared to the general population sample cohort (provided by Fitbit), a linear regression model was fit for each sleep variable:

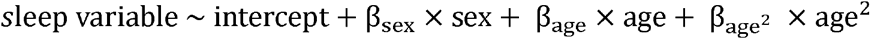

where the intercept was approximately the mean of the sleep variable across all users in the cohort of interest. Sex was defined as 1=Male, 2=Female; age was defined in years. Age was included as a polynomial, given known non-linear relationships with many sleep metrics with age.^24^ The standard errors of each of the coefficients were calculated and the significance of the intercept (mean) of each sleep variable for the epilepsy cohort was compared to the general population cohort using a t-statistic test with unequal variances.

### Sleep duration prior to seizure-free and seizure days

To evaluate the effect of sleep duration on seizure risk, we considered both day-to-day sleep duration on seizure risk and multi-day patterns of sleep duration leading up to seizures. For the day-to-day analyses, sleep durations in pre-seizure windows were compared to non-seizure windows using the Wilcoxon rank-sum test to assess the null hypotheses that two sets of measurements (seizure vs seizure-free) were drawn from the same distributions. For each participant, a pre-seizure window of 3 days or the baseline seizure frequency (whichever was lower) was considered. For instance, for participants with an average seizure frequency of one every 2.5 days, only 2-day pre-seizure windows were used. To adjust for multiple comparisons, the p-value threshold was set at 0.05 / (number of days analysed).

The effect of multi-day sleep duration patterns (“multi-day analyses”) prior to seizure days were assessed by comparing a straight line (i.e., slope of zero) to the slope of the average sleep duration pattern in the 3 days leading up to seizure days. Only participants with an average seizure frequency of less than 3 seizures per week were considered for the multi-day analysis. The slope of the average sleep duration was calculated using a linear least squares regression line. The significance of the sleep duration slope was assessed using a t-statistic test (herein referred to as the “slopes test”).^25^

Note that only the lead seizure after each pre-seizure window was considered. For nocturnal events, the night prior to the night of the nocturnal event was defined as the pre-seizure sleep.

### Oversleep and undersleep prior to seizure days

To evaluate the effect of oversleep and undersleep, we used a mixed effects logistic regression model with random subject-specific intercept (consistent with previous literature)^11,12^. For each patient, days were categorised into decreased, baseline, and increased sleep duration. Baseline sleep was defined as sleep durations falling within the interquartile range (≥25th percentile and ≤75th percentile). Durations below the 25th percentile and above the 75th percentile were classed as decreased and increased sleep duration, respectively. Alteration of sleep duration was assumed to play a role if a seizure occurred within 48 h after an increase or decrease in sleep duration. Each patient contributed multiple days of observation into each model, therefore the random subject-specific intercept was used to take within-subject correlation into account. The sleep categories (decreased, baseline, and increased) were the fixed effects.

We also considered the effect of oversleep, undersleep and nocturnal seizures during different seizure risk states (low, baseline and high risk). These risk states were determined retrospectively for each seizure using a seizure-cycles based algorithm, similar to previous work.^26^ In contrast to Karoly et al.,^26^ we selected the two strongest cycles (using SI values) between 4 and 70 days to forecast seizure risk (i.e., we did not consider cycles less than 4 days).

### Sleep onset and offset prior to seizure-free and seizure days

To evaluate the effect of bedtime (sleep onset) and wake time (sleep offset), the Wilcoxon rank-sum test was used to assess the null hypotheses that two sets of measurements (seizure vs seizure-free days) were drawn from the same distributions (p < 0.05). For nocturnal seizures, we defined bedtime as the sleep onset time on the night of the seizure, and waketime as the sleep offset time on the night before the night of the seizure.

## Data Availability

Excluding participants who did not consent to share their data publicly, deidentified data are available on Figshare (as DOI 10.6084/m9.figshare.20381298).

## Results

### Sleep in epilepsy compared to the general population

Participants who had at least 28 nights of sleep recorded with their smartwatch (N = 44) were included. A total of 3894 self-reported seizures (M = 88, SD = 130, Range = 1 - 523) and 17078 nights (M = 388, SD = 351, Range = 32-1701) of recorded sleep were included in the study. See Supplementary Table 1 for detailed participant demographic information. Figure 1 shows an example of two consecutive nights of sleep recorded using the smartwatch.

**Figure 1.**
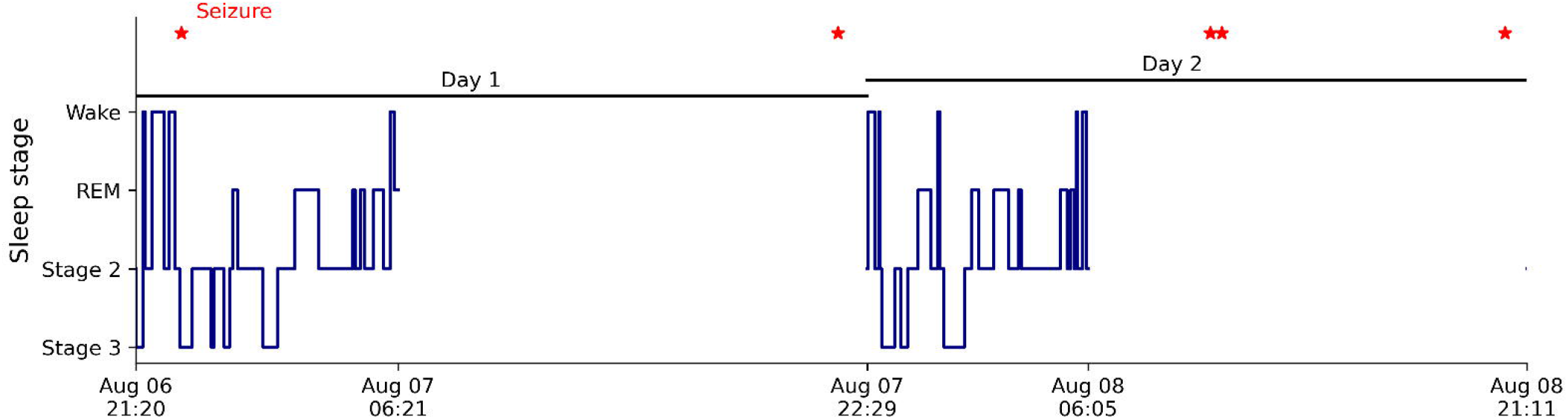
Example sleep hypnogram of two consecutive nights of sleep recorded using the smartwatch. Seizures are indicated by red stars.

Data provided by Fitbit were used to determine sleep variable averages of a representative sample of 37921 Fitbit users (46.74+/-14.91 years; range 15-85 years; 69.7% female), each with 28 consecutive nights of Fitbit sleep data. Figure 2 shows the results of linear model coefficients (and their standard errors) for mean, age and sex parameters of the epilepsy cohort compared to the general population sample. There was a significant difference in the percentage of duration spent awake during overnight sleep (i.e., wake after sleep onset, WASO) for users with epilepsy compared to the general population sample (p < 0.05 using t-test). There was also a trend for users with epilepsy to sleep longer overnight and spend more time in deep sleep compared to users in the general population sample (total sleep duration: 9.08 ± 1.75 vs 7.05 ± 0.04 and Deep minutes: 124.15 ± 24.48 vs 87.16 ± 0.68) (Supplementary Figure 1), although these differences were not significant. See Supplementary Table 2 for a breakdown of the residual standard errors for each sleep variable (a measure of the fit of the linear regression model).

**Figure 2.**
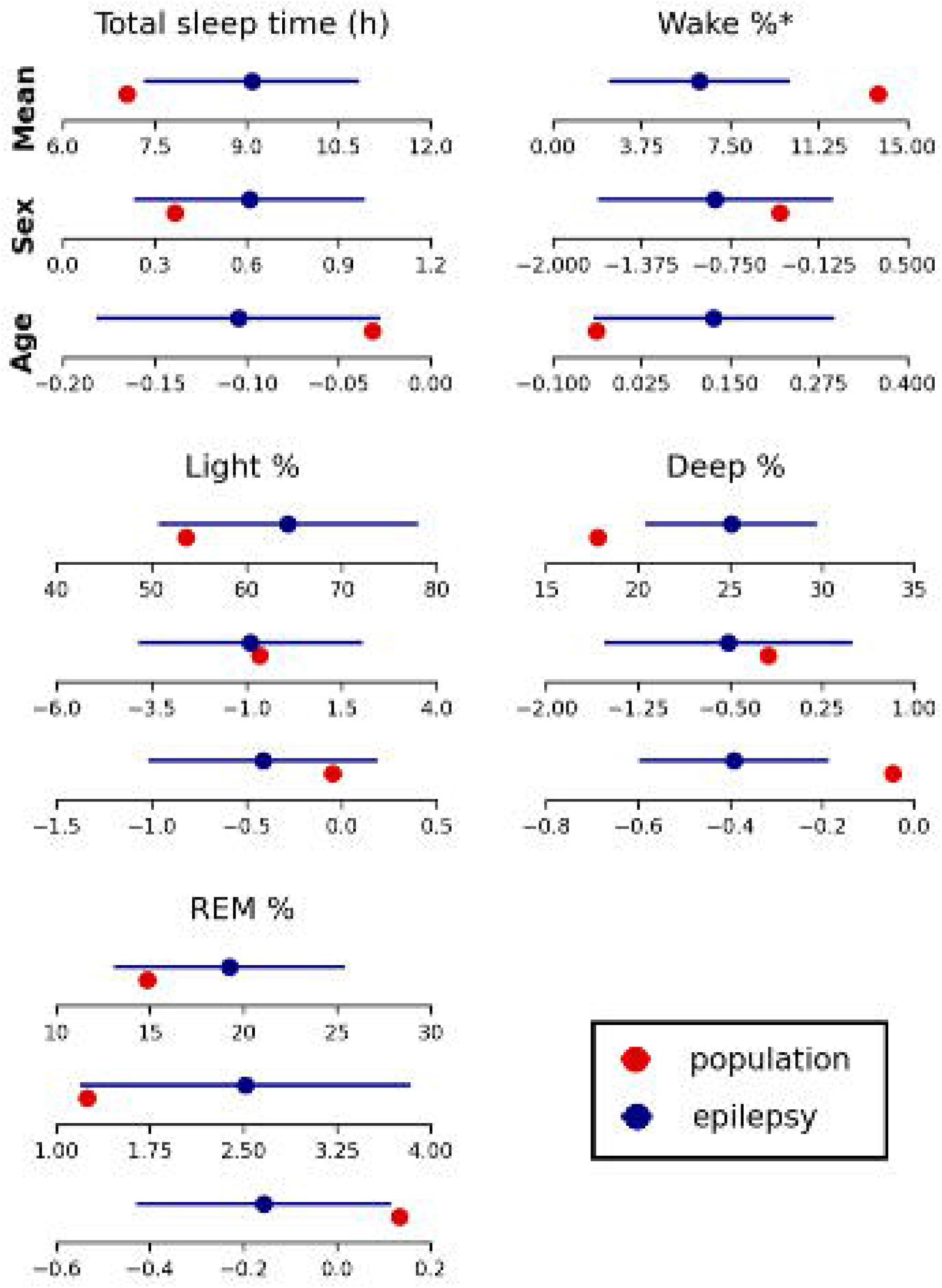
Linear regression coefficients of average time sleeping overnight, and proportions of time spent in wake/light/deep/REM sleep states across a general population sample (red) compared to the epilepsy cohort (navy). Changes in average values are also shown with differing sex and age. Sex is females relative to males, so a negative value indicates females have lower amounts of the sleep variable of interest compared to males. Age is in years so negative values indicate that ageing results in lower amounts of the sleep variable of interest. Error bars show standard errors of the linear regression parameters for each sleep variable. *Indicates significant difference between sleep variable mean of the epilepsy cohort compared to the general population.

The remaining results sections includes participants with epilepsy who reported at least 10 seizure days following a night that was recorded on their smartwatch (N = 29).

### Sleep duration prior to seizure-free and seizure days

When considering sleep duration, day-to-day and multi-day sleep distributions were investigated. In day-to-day sleep analysis, distribution of sleep duration on day 1 to day 3 leading up to seizure and seizure-free days were compared (Figure 3). Only 2 out of 29 participants had significant differences between their day-to-day sleep durations on at least one of the three days preceding a seizure or seizure-free day (relative changes and p-values are provided in Supplementary Table 3). In both participants, the significant difference in sleep duration was an increase in their pre-seizure sleep (relative to their seizure-free sleep). In one of these cases (P9), the increase was detected on the day before the seizure (Supplementary Figure 2).

**Figure 3.**
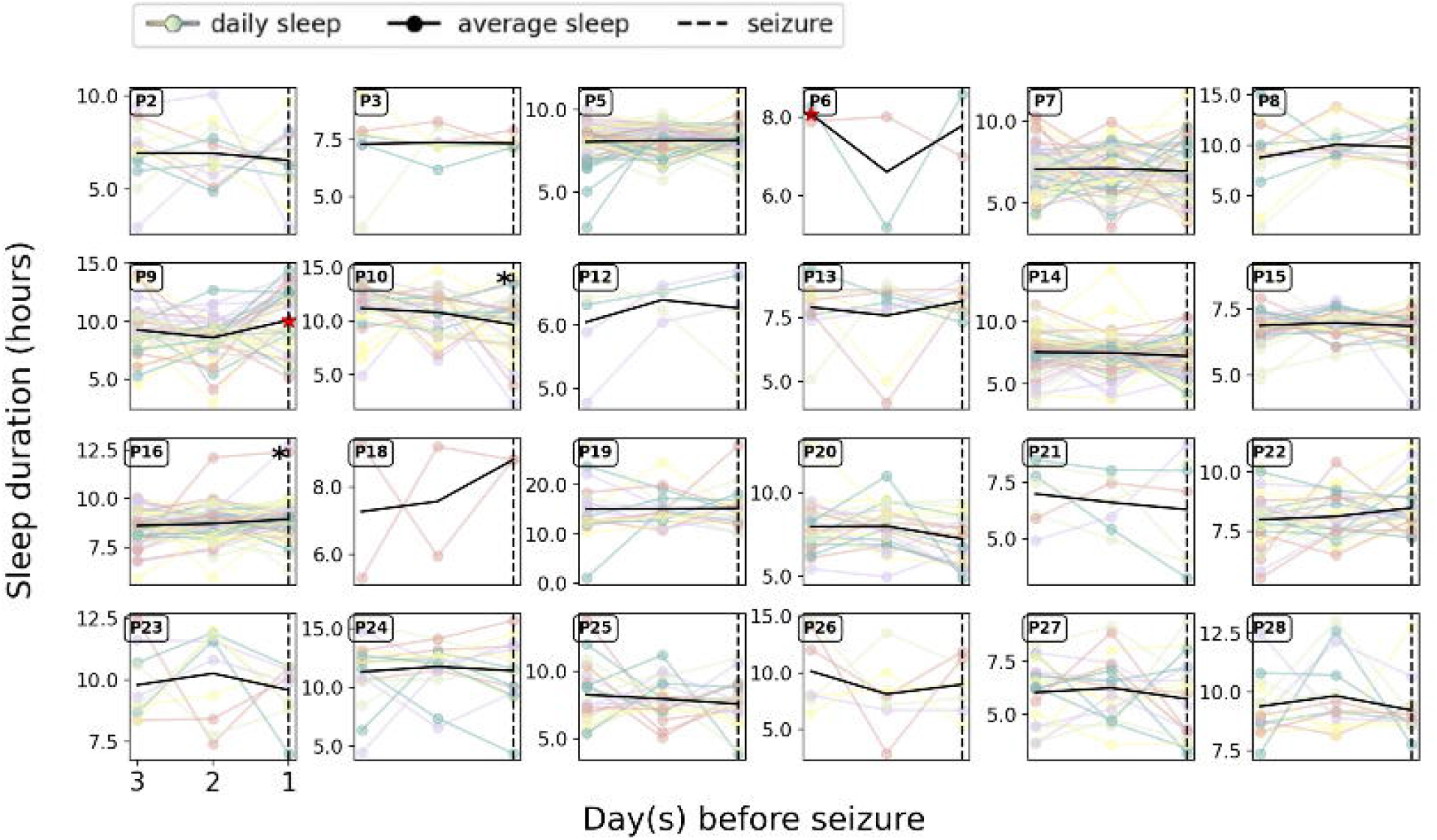
Sleep duration trends for three days prior to seizure days. For each subpanel, coloured lines represent sleep duration for the three days prior to an individual seizure, and black lines indicate the average pre-seizure sleep duration (i.e., the mean of all coloured lines). *red asterisks indicate a significant difference (using the Wilcoxon ranksum test with Bonferroni correction) between sleep duration distributions on the nth-day preceding seizure and seizure-free days. *black asterisks indicate significant (p < 0.05) slope of 3-day sleep duration trend prior to a seizure day.

For multi-day sleep analysis, a window of three days of average sleep duration prior to seizure days were plotted (Figure 3) and the slopes test was utilised to determine whether a significant trend was present in the average sleep durations leading up to seizure days. Two out of 24 participants (P10 and P16) displayed a significant trend in their multi-day sleep duration slopes prior to seizure days. For P10 there was a decrease in their average sleep duration in the week leading up to a seizure; conversely, P16 showed an increase in average sleep duration in the week leading up to a seizure. On the whole, Figure 3 demonstrates high inter-seizure variability in participants’ pre-seizure sleep durations (coloured lines). However, on average, changes in sleep duration (black lines) were not consistent across seizures, and pre-seizure sleep duration changes were not significant for the majority of individuals.

### Oversleep and undersleep prior to seizure days

To assess the interaction between day-to-day deviations in sleep duration and seizure probability, we used logistic regression with a random subject-specific intercept (consistent with Dell et al.)^11^. For each patient, days were categorised into decreased, baseline, and increased sleep duration. For most (18/29) participants, sleeping for longer durations was correlated with increased seizure likelihood in the following 48h (Figure 4). At the group level, a logistic regression model revealed that when patients slept longer than the 75th percentile, there was a 20% increase in the odds of a seizure in the following 48 h when compared to baseline (OR=1.20, CI = [1.07, 1.35], p = 0.001). For our cohort, the 75th percentile equates to an average of 8.24 (±0.60) h of sleep, which is 0.79 (±0.35) h longer than their median sleep duration. Sleeping less than the 25th percentile (6.69 ± 0.78 h), did not have any significant group effect on the odds of a seizure (p=0.516).

**Figure 4.**
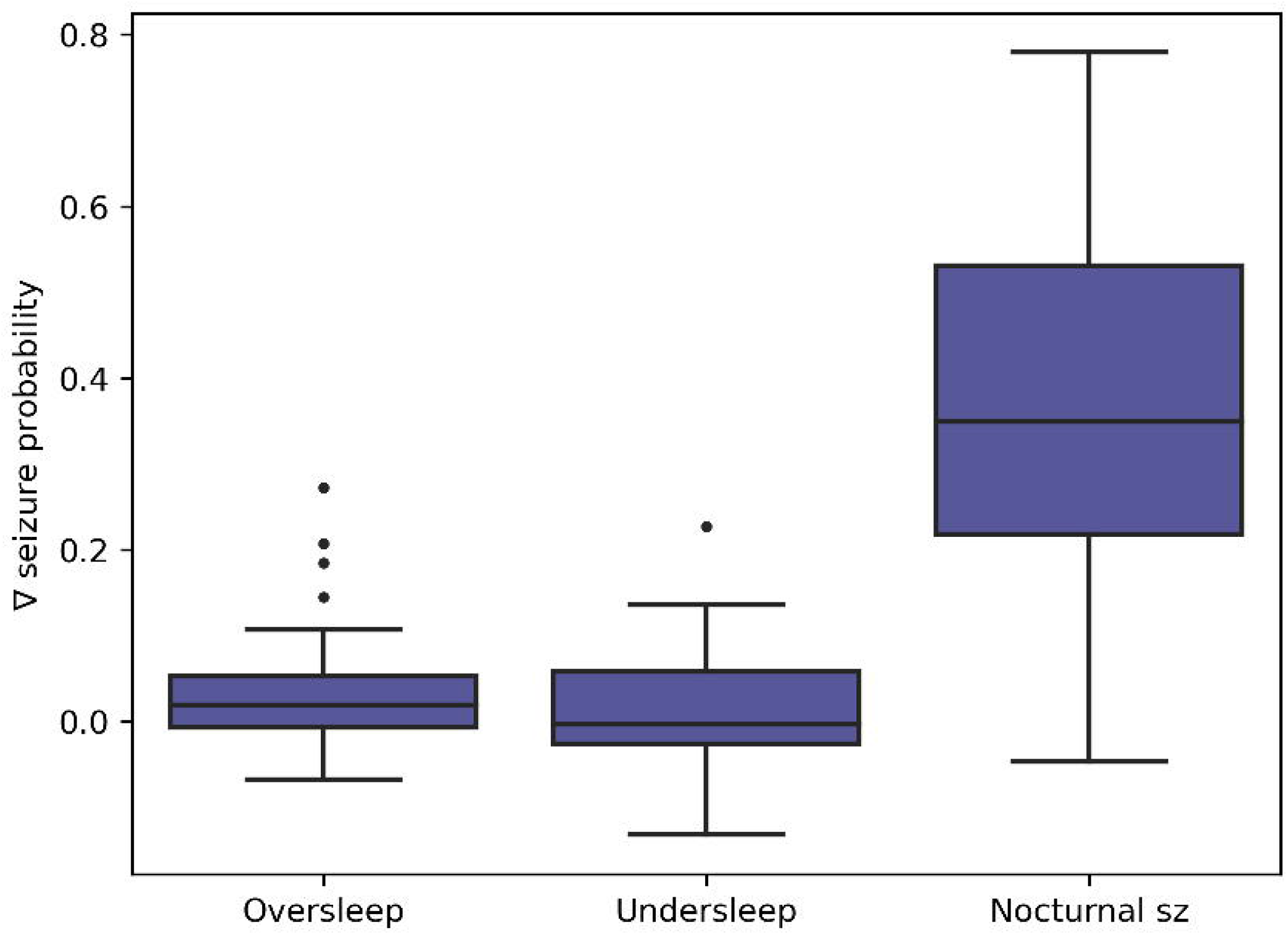
Change in seizure risk relative to baseline conditions across participants. Positive values reflect increased risk, negative values reflect decreased risk compared to the baseline condition. For each participant, nights of oversleep and undersleep were defined as being greater than the 75^th^ percentile or less than the 25^th^ percentile of sleep duration, respectively (with baseline sleep duration defined to be between the 25^th^ and 75^th^ percentiles). The change in seizure risk following nocturnal seizures was only analysed for the 21 participants who reported nocturnal events, with the baseline condition including all nights without reported nocturnal events.

Notably, almost all individuals with reported nocturnal events (19/21) (i.e., events that occurred during overnight sleep) had increased risk of seizures in the following 48h (Figure 4). Nocturnal events were also associated with increased median sleep duration (8.23h compared to 7.64h, p < 0.001 using the Wilcoxon ranksum test). Therefore, the association between oversleep and seizure occurrence can most likely be attributed to nocturnal events increasing both sleep duration and subsequent seizure risk. Nevertheless, oversleep remained significantly associated with seizure risk when nocturnal events were included as a covariate in the logistic regression model, perhaps due to unreported nocturnal seizures driving longer sleep duration.

The risk level of each seizure was also considered in the context of oversleep, undersleep and nocturnal events (Figure 5). Sleeping longer than the 75^th^ percentile was only significantly correlated with increased seizure likelihood in the following 48h if the seizure occurred during high-risk state. Interestingly, oversleep did not significantly correlate with seizure likelihood in the following 48h when the participant was in a low-or baseline-risk state. Sleeping less than the 25^th^ percentile did not have any significant effect on the odds of a seizure in any risk state. However, nocturnal events significantly increased the risk of seizures in the following 48h in all risk states (low, baseline and high), although visually the risk was greater if the seizure occurred in a high-risk state.

**Figure 5.**
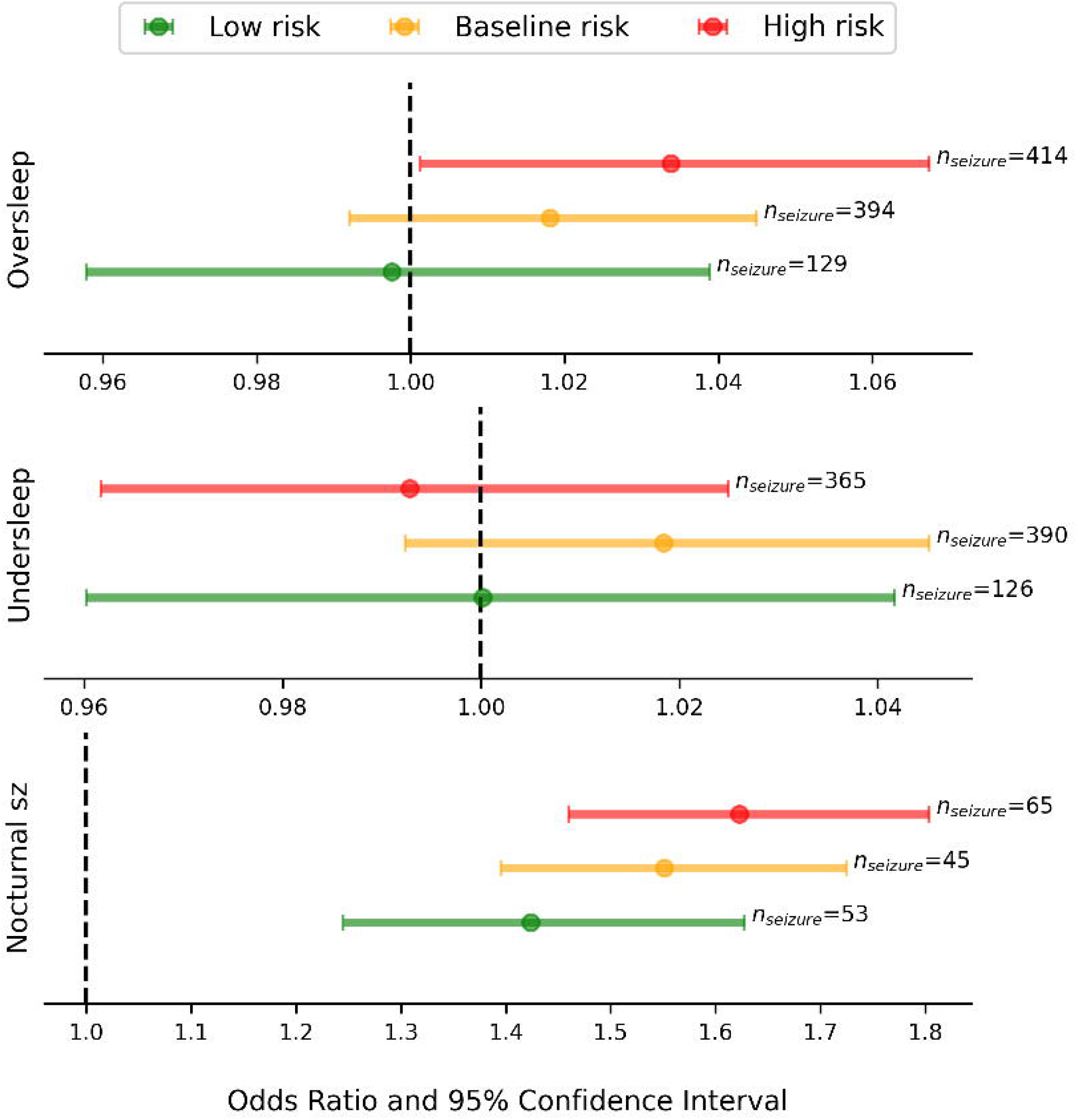
Odds ratios of a seizure (in high, baseline or low risk) occurring within 48 hours of an oversleep night, an undersleep night or a nocturnal seizure. Coloured dots show odds ratios and bars show 95% confidence intervals. Positive values reflect increased risk, negative values reflect decreased risk compared to the baseline sleep condition. For each participant, nights of oversleep and undersleep were defined as being greater than the 75^th^ percentile or less than the 25^th^ percentile of sleep duration, respectively (with baseline sleep duration defined to be between the 25^th^ and 75^th^ percentiles). Nocturnal seizures were only analysed for the 21 participants who reported nocturnal events, with the baseline condition including all nights without reported nocturnal events. n_seizure_ indicates the number of seizures that occurred in that state across the cohort.

### Sleep onset and offset on seizure-free and seizure days

For 11 of 29 subjects, there was a significant difference between either sleep onset/offset (bed/wake) on seizure days compared to seizure-free days (see Supplementary Table 4). Four people had a difference in their waketimes only (one earlier than normal, three later than normal), two people had a difference in their bedtimes only (one earlier than normal, one later than normal) and five had changes in both bed and wake times on seizure days compared to seizure-free days. Figure 6 highlights three examples of study participants: P9 woke significantly later prior to seizure days, P18 slept significantly earlier prior to seizure days and P20 slept and woke significantly later prior to seizure days, compared to seizure-free days. Interestingly, P20 previously appeared to have no difference in sleep duration prior to seizure-free and seizure days (Supplementary Figure 2 and Supplementary Table 3).

**Figure 6.**
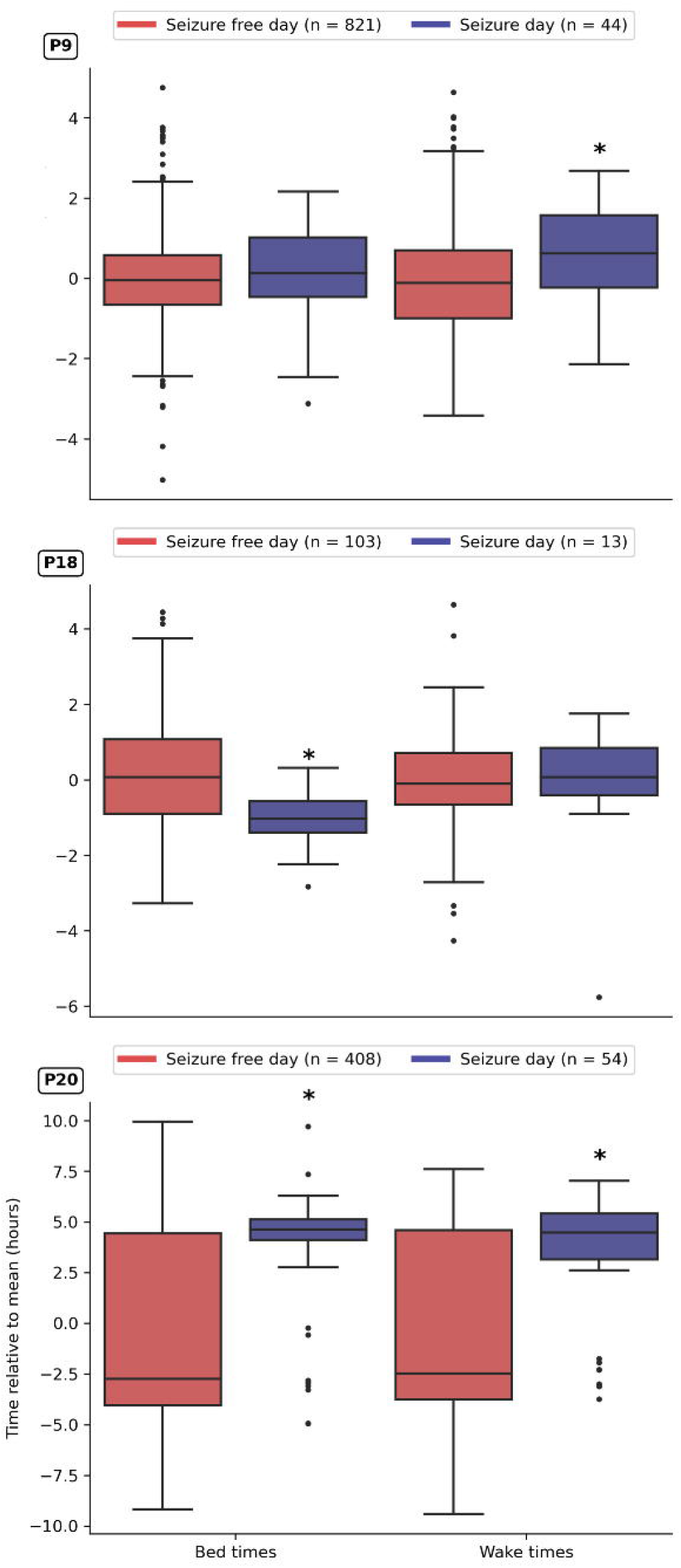
Box plot distributions of bed- and wake-times on seizure-free days (red) compared to seizure days (navy) for P9, P18 and P20. *indicates a significant difference between bed or wake time distributions of seizure-free days compared to seizure days.

Note that only two of the participants (P9 and P26) across the cohort had a significant predominance of seizures on a particular day of the week (Supplementary Figure 3), reducing the likelihood that seizure risk was mediated by confounding weekday effects, rather than sleep onset/offset times. We did not investigate the effect of shift work on seizure risk in this study.

## Discussion

Despite significant interest in the association between sleep and seizure risk, there is a notable lack of long-term quantitative studies, with just one prior study using invasively recorded EEG^11^ and three studies of self-reported sleep and seizure diaries.^12,13,27^ This study is the first to examine long-term associations between sleep and self-reported seizures using non-invasive wearables. Subjects wore smartwatch devices for an average of 13 months while undergoing their normal day-to-day activities, representing the largest objective analysis of sleep and subsequent seizure occurrence.

The current study was the first to compare sleep in epilepsy to the general population in an at-home setting. Sleep duration and composition (proportion of light, deep and REM) in the epilepsy cohort were compared to a large representative population of more than 37,000 Fitbit users (Figure 2). This large population sample provided an unprecedented ability to identify differences in sleep patterns for people with epilepsy, based on directly comparable recordings (i.e., from a Fitbit). The only significant difference was for wake after sleep onset duration as a proportion of total sleep duration: people with epilepsy spent significantly less time awake during sleep than the general population. In addition, people with epilepsy spent more time in deep sleep and slept an average of 2 hours longer than the general population (although these trends were not significant, due to higher individual variability). There was no noticeable difference in REM sleep duration or proportion between epilepsy and general populations. Conversely, a recent study found a decreased proportion of REM sleep in people with epilepsy compared to healthy controls,^28^ although sleep data were collected from an inpatient setting, so were unlikely to reflect normal sleeping patterns.

Both the general population and epilepsy cohort models showed women slept longer than men, and sleep duration decreased with age (Figure 2). Longer sleep durations for people with epilepsy may result from numerous factors, including epileptologists suggesting their patients prioritise sleep (Rossi et al., 2020), sleeping longer after seizures,^11^ adverse effects of anti-seizure medications, including somnolence and fatigue,^29^ and the higher likelihood of coexisting sleep conditions that reduce sleep quality, such as obstructive sleep apnea.^30^

Figure 4 illustrates that increased sleep durations and oversleep were weakly associated with increased seizure likelihood. This finding was contrary to previous studies showing that increased sleep duration reduced the risk of seizures in the following 48 hours for both electrographic^11^ and self-reported^12^ seizures. However, one other previous study^27^ found no significant effects of sleep duration and seizure occurrence. It is important to note that these findings address everyday fluctuations in sleep duration and may not accurately reflect extreme scenarios, such as the effects of substantial, forced sleep deprivation on seizure risk. In the current study, the link between oversleep and seizure risk was primarily driven by the occurrence of reported nocturnal seizures (Figure 4). Nocturnal seizures were associated with significantly longer sleep durations (consistent with Dell et al.)^11^ and increased the risk of subsequent seizure occurrence. Interestingly, oversleep still showed independent predictive value in addition to nocturnal seizure occurrence, potentially due to the presence of unreported nocturnal events or the daytime reporting bias (where nocturnal events are inaccurately reported as occurring after sleep offset). However, the association between oversleeping and increased seizure risk in the following 48h may also be related to anti-seizure medication adherence, whereby patients forget to take their medication or delay their usual medication time.

Interestingly, in the present work, correlation between oversleep and increased seizure likelihood was only detected during the high-risk state (Figure 5). This suggests that sleep duration (specifically sleeping longer than normal) influences seizure risk only if the endogenous seizure risk rhythm is in a high-risk state when the seizure occurs. This result is in line with a stochastic, multi-stable dynamical model of seizure transitions, whereby the brain state approaches a critical transition point leading to increased susceptibility for seizures triggered by either noisy perturbations or steady parameter change.^31^ In other words, during high-risk periods, when the brain is already closer to a tipping point, oversleeping further reduces the critical barrier or ‘seizure threshold’, although oversleep by itself may be insufficient to trigger seizures. Nocturnal seizures were also correlated with seizure risk in the following 48 hours irrespective of the endogenous seizure risk cycle, suggesting another mechanistic driver such as seizure clustering; however, being in a high-risk state seemed to exacerbate the risk associated with nocturnal events, compared to a baseline-or low-risk state (Figure 5).

Fluctuations in sleep onset and offset times were significantly associated with seizure risk for more participants (11 out of 29) than changes in sleep duration (4 out of 29 had either a multiday or day-to-day trend). Notably, two of the individuals whose sleep duration related to seizure occurrence also showed significant changes in their bed or wake times prior to seizures. These findings suggest that monitoring sleep onset and offset is more informative than duration. For instance, later sleep onset and offset could be significantly associated with seizure risk (e.g., participant P20) without altering sleep duration. Conversely, earlier sleep onset and offset could also be associated with seizure risk without noticeable changes in duration (e.g., participant P28). Overall, the effect of sleep onset/offset times on seizure risk were highly subject specific, with examples of both later and earlier bed or wake times associated with seizure risk (Figure 6 and Supplementary Table 4). This suggest that, in most cases, maintaining a consistent sleep schedule may help reduce seizure frequency. It is not clear why maintaining a consistent sleep schedule reduces seizure risk, but it may be related to the inhibitory effect of REM sleep (which is impacted by sleep timing) or anti-seizure medication adherence. Nonetheless, these findings highlight the importance of taking an individual-specific approach to sleep counselling in epilepsy management and reiterate the value of wearables in epilepsy research, given that wearable devices typically perform well at detecting sleep onset and offset.^32,33^

This study did not focus on sleep composition due to limitations in the wearable sleep staging algorithm for some people^22,23,34^, although some subjects did appear to have differences in their sleep composition prior to seizure days compared to seizure-free days (Supplementary Figure 4 and Supplementary Table 5). Differences in sleep composition prior to seizures were highly individual-specific and likely reflected changes in sleep duration in many cases. Similarly, Dell et al.^11^ observed no significant group-level changes in sleep composition prior to electrographic seizure occurrence.

This study had several limitations. Self-reported seizures have known drawbacks including their potential for inaccuracy and bias towards daytime seizures and seizures of higher severity or impaired awareness.^35^ However, seizure diaries remain an important source of information and provide sufficiently accurate data for many applications.^36,37^ With long-term data, seizure diaries may reliably detect underlying patterns even in the presence of signal noise.^5,26,37,38^ Furthermore, many of the existing assumptions regarding sleep and seizure risk are based on sleep and seizure diary accounts.

Results in this study were based on sleep parameters detected using wearable smartwatch devices (Fitbit), particularly total sleep duration, sleep onset and offset, and classification of sleep stages. Recent studies have shown that newer Fitbit devices, which rely on multiple signals to detect sleep stages, have satisfactory performance with regards to their total sleep duration estimations and the transition from wake to sleep and sleep to wake.^32,33^ However, for classification of sleep stages, wearable smartwatches are not yet suitable substitutes for polysomnography,^22,23,34^ with a few exceptions in their ability to detect transitioning from certain states (deep sleep to wake state, and light sleep to REM sleep) and the likelihood of remaining in REM sleep.^34^ Despite this lack of confidence in classification of sleep stages, it is still possible to detect clear individual-specific relationships between seizures and other sleep parameters, notably total sleep duration, sleep onset and offset, and oversleep and undersleep, which indirectly relate to sleep architecture and sleep quality. Moreover, wearable sleep monitors provide an objective account of sleep quality, compared to most prior long-term sleep and epilepsy studies that use sleep diaries.^12,13,27^

## Conclusion

The current study found no strong evidence that moderately less sleep (i.e., 2-3 hours less than average) increased the likelihood of seizures, consistent with previous work. On the other hand, contrary to previous studies, the current results showed oversleep was weakly associated with increased, rather than decreased, seizure risk. This seemingly contradictory finding was most likely due to nocturnal seizures driving longer sleep durations or a medication adherence covariate. This study also demonstrated that fluctuations in bedtime and waketime were more informative than sleep duration for identifying seizure risk. The presented results have important clinical implications and can guide sleep counselling in clinical epilepsy management. For instance, the current work suggested it is more important for people with epilepsy to maintain a consistent sleep routine (bed and wake times) than to maximise their sleep duration. However, the relationship between bed and wake times was also highly individual-specific, underscoring the potential utility for wearable devices to personalise sleep and seizure management plans.

## Data Availability

Excluding participants who did not consent to share their data publicly, deidentified data are available on Figshare (as DOI 10.6084/m9.figshare.20381298, to be made available upon publication).

https://figshare.com/articles/dataset/Sleep_and_seizures_in_epilepsy/20381298

## Abbreviations

IEDs: interictal epileptiform discharges

## Acknowledgements

The authors would like to thank our collaborators within the Clinical Health Research Team at Fitbit and Fitbit users who are willing to share their data to develop a better understanding of real-world sleep patterns.

## Supplementary Material

Supplementary material is available at *Brain* online.

