## Supplement for "Sleep and seizure risk in epilepsy: Bed and wake times are more important than sleep duration"

### Supplementary Table 1. Participant demographic information.

Epilepsy types are separated into Focal, Multi-focal, GGE or Focal and Generalised. The epilepsy syndrome or lobar epileptogenic zone is presented in brackets where the data is available. (T = temporal, TO = temporo-occipital, TP = temporo-parietal, HH = hypothalamic hamartoma, DEE = Developmental and Epileptic Encephalopathy, JME=juvenile myoclonic epilepsy, JAE=juvenile absence epilepsy, LGS=Lennox-Gastaut syndrome, GGE=genetic generalised epilepsy).

| Participant | Age range | Gender | Epilepsy Syndrome | Anti-Seizure Medication | Sleep days | Number of seizures |
|---|---|---|---|---|---|---|
| P1 | 26-30 | F | GGE (JAE) | Sodium Valproate, Lamotrigine, Perampanel | 171 | 171 |
| P2 | 31-35 | M | Focal (T) | Carbamazepine | 360 | 38 |
| P3 | 41-45 | F | | | 205 | 59 |
| P4 | 31-35 | F | LGS | | 88 | 523 |
| P5 | 26-30 | F | Multi-focal | Oxcarbazepine | 676 | 143 |
| P6 | 31-35 | M | | | 68 | 11 |
| P7 | 36-40 | F | Focal (T) | Zonisamide, Lamotrigine, Clonazapam, Medicinal Cannabis | 1695 | 518 |
| P8 | 56-60 | M | Focal | Sodium Valproate, Lamotrigine, Perampanel | 135 | 23 |
| P9 | 31-35 | F | Focal | Levetiracetam, Lacosamide | 865 | 44 |
| P10 | 31-35 | F | Multi-focal | None | 400 | 256 |
| P11 | 36-40 | F | | | 303 | 158 |
| P12 | 41-45 | F | Focal | Levetiracetam, Topiramate | 412 | 13 |
| P13 | 66-70 | F | | | 80 | 37 |
| P14 | 36-40 | M | Focal and Generalized (DEE) | Lamotrigine, Clonazepam, Oxcarbazepine, Zonisamide | 465 | 286 |
| P15 | 66-70 | M | Focal | Brivaracetam, Levetiracetam | 1701 | 80 |
| P16 | 31-35 | F | Focal (T) | Lacosamide, Brivaracetam, Zonisamide, Losartan | 695 | 147 |
| P17 | 26-30 | F | Focal (T) | Levetiracetam, Topiramate, Lacosamide | 601 | 341 |
| P18 | 41-45 | F | | | 116 | 76 |
| P19 | 36-40 | M | Focal (TP) | Levetiracetam, Topiramate, Lamotrigine | 196 | 52 |
| P20 | 26-30 | M | Focal (TO) | Levetiracetam, Lamotrigine | 463 | 113 |
| P21 | 31-35 | F | Focal (T) | Zonisamide, Lacosamide | 254 | 38 |
| P22 | 56-60 | F | Focal (T) | Topiramate, Lamotrigine, Carbamazepine | 575 | 31 |
| P23 | 51-55 | F | Multi-focal | Sodium Valproate, Pregabalin, Lacosamide | 449 | 27 |
| P24 | 71-75 | M | Focal (T) | None | 700 | 30 |
| P25 | 26-30 | F | Focal (F) | Clobazam, Lacosamide, Levetiracetam, Oxcarbazepine | 292 | 250 |
| P26 | 66-70 | F | GGE (JAE) | Sodium Valproate, Lamotrigine | 118 | 13 |
| P27 | 56-60 | M | Focal (T) | Levetiracetam, Topiramate, Lamotrigine | 425 | 213 |
| P28 | 21-25 | F | Focal (T) | Levetiracetam, Topiramate, Lacosamide | 556 | 118 |

| | | | | | | |
|---|---|---|---|---|---|---|
| P29 | 26-30 | M | | None | 159 | 85 |
| P30 | 51-55 | F | Multi-focal | Brivaracetam, Phenobarbital, Clonazepam, Carbamazepine | 284 | 0 |
| P31 | 51-55 | F | Focal (HH) | Lacosamide, Zonisamide, Clobazam | 379 | 0 |
| P32 | 56-60 | F | | | 74 | 0 |
| P33 | 46-50 | M | | | 245 | 0 |
| P34 | 26-30 | M | GGE (JME) | Sodium Valproate, Pregabalin, Lacosamide | 210 | 0 |
| P35 | 26-30 | F | Focal (T) | Sodium Valproate | 581 | 0 |
| P36 | 16-20 | F | | | 168 | 0 |
| P37 | 46-50 | F | Focal (HH) | Lacosamide, Zonisamide, Clobazam | 432 | 0 |
| P38 | 36-40 | F | | | 205 | 0 |
| P39 | 36-40 | M | | | 32 | 0 |
| P40 | 31-35 | F | GGE | Carbamazepine, Clobazam | 565 | 0 |
| P41 | 31-35 | F | GGE (JME) | Sodium Valproate | 231 | 0 |
| P42 | 21-25 | F | Focal (T) | Brivaracetam, Lacosamide, Perampanel, Lamotrigine | 275 | 0 |
| P43 | 46-50 | F | | | 57 | 0 |
| P44 | 21-25 | F | | | 117 | 0 |
| | **M = 40.3 SD = 14.0** | **31 Female 13 Male** | | | **M = 388 SD = 351** | **M = 89 SD = 130** |

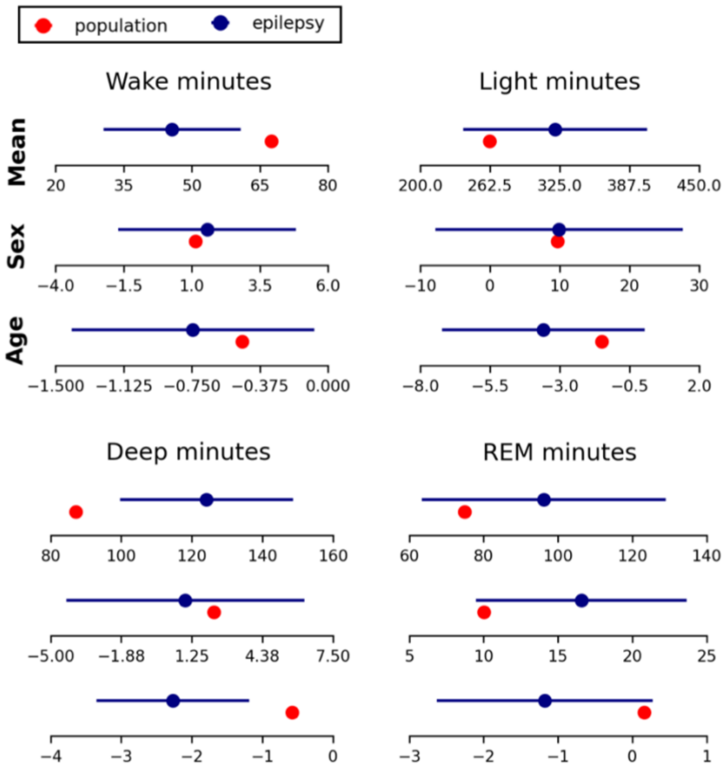

**Supplementary Figure 1. Linear regression coefficients of average duration spent in wake/light/deep/REM sleep across a general population sample (red) compared to the epilepsy cohort (navy).** Changes in average values are also shown with differing sex and age. Sex is females relative to males, so a negative value indicates females get less of the sleep variable of interest compared to males. Error bars show standard errors of the linear regression parameters for each sleep variable. *Indicates significant difference between sleep variable mean of the epilepsy cohort compared to the general population.

**Supplementary Table 2.** Residual standard errors of sleep variable linear regression models for the epilepsy cohort and the general population sample.

| | Residual Standard Error | |
|---|---|---|
| | **General Population Sample** | **Epilepsy** |
| **Total sleep** | 0.7187 | 1.1284 |
| **Wake minutes** | 12.3865 | 9.7434 |
| **Wake %** | 1.8455 | 2.4754 |
| **Light minutes** | 35.6115 | 53.0283 |
| **Light %** | 4.3869 | 8.7890 |
| **Deep minutes** | 13.2567 | 15.7540 |
| **Deep %** | 2.7282 | 3.0230 |
| **REM minutes** | 18.0831 | 21.1506 |
| **REM %** | 3.4081 | 3.9770 |

**Supplementary Table 3. Average daily sleep durations before seizure days, relative to seizure-free days.** *indicates significant difference between average sleep durations on nth day before seizure and seizure-free days.

| Participant | Day 1 | Day 2 | Day 3 |
|---|---|---|---|
| **P1** | +0.25 (0.9206) | -0.33 (0.4931) | |
| **P2** | -0.63 (0.0448) | -0.59 (0.0592) | -0.1 (0.1321) |
| **P3** | -0.05 (0.905) | -0.0 (0.8059) | +0.13 (0.5624) |
| **P4** | -0.22 (0.897) | +0.34 (0.8171) | |
| **P5** | -0.04 (0.0885) | -0.07 (0.0808) | -0.09 (0.0731) |
| **P6** | +0.21 (0.0174) | -0.79 (0.0272) | **+0.45 (0.0149*)** |
| **P7** | -0.49 (0.0964) | -0.08 (0.4732) | -0.13 (0.311) |
| **P8** | +0.43 (0.6441) | +0.27 (0.319) | -0.02 (0.5575) |
| **P9** | **+0.43 (0.0032*)** | -0.01 (0.5798) | +0.16 (0.3086) |
| **P10** | -0.56 (0.2005) | -0.02 (0.7635) | +0.14 (0.5846) |
| **P11** | +0.17 (0.5346) | +0.36 (0.5436) | |
| **P12** | -0.31 (0.9215) | -0.05 (0.8203) | -0.27 (0.9499) |
| **P13** | +0.08 (0.2252) | +0.08 (0.6182) | +0.22 (0.3108) |
| **P14** | -0.1 (0.2826) | -0.04 (0.5263) | +0.05 (0.6489) |
| **P15** | +0.08 (0.6747) | +0.1 (0.0999) | +0.07 (0.6916) |
| **P16** | +0.17 (0.0771) | +0.06 (0.9978) | -0.03 (0.5772) |
| **P17** | +0.12 (0.0906) | -0.11 (0.9039) | |
| **P18** | +2.37 (0.1975) | +0.94 (0.1975) | +0.49 (0.3604) |
| **P19** | +0.25 (0.2208) | +0.44 (0.2395) | +0.12 (0.1896) |

| | | | |
|---|---|---|---|
| **P20** | -0.07 (0.3068) | +0.54 (0.1786) | +0.33 (0.3506) |
| **P21** | -0.31 (0.4116) | -0.69 (0.6909) | -0.06 (0.4597) |
| **P22** | +0.03 (0.4463) | -0.15 (0.4093) | +0.08 (0.5291) |
| **P23** | +0.46 (0.9336) | +1.44 (0.3817) | -0.2 (0.8678) |
| **P24** | -0.1 (0.6106) | +0.24 (0.2011) | +0.2 (0.5934) |
| **P25** | +0.08 (0.6908) | +0.29 (0.6067) | +0.2 (0.5007) |
| **P26** | +0.31 (0.4204) | -0.25 (0.8057) | -0.33 (0.1664) |
| **P27** | -0.76 (0.1465) | -0.59 (0.4635) | -0.59 (0.2699) |
| **P28** | -0.25 (0.964) | +0.11 (0.275) | -0.05 (0.578) |
| **P29** | -0.23 (0.6859) | +0.09 (0.9648) | |

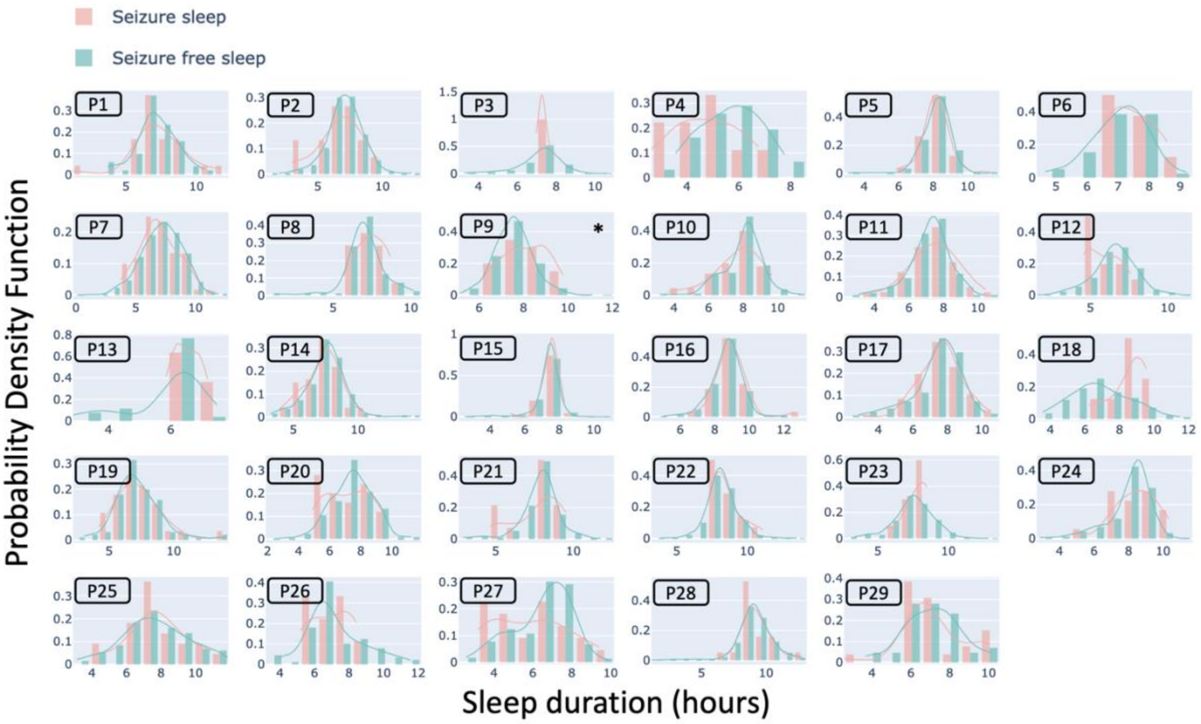


**Supplementary Figure 2. Distribution of sleep duration (hours) the day preceding seizure (orange) and seizure-free (green) days across the cohort.** *Indicates significant difference on sleep distributions preceding seizure days compared to seizure-free days.

**Supplementary Table 4. Bedtimes (relative to the mean bedtime) on seizure days compared to seizure-free days and waketimes (relative to the mean waketime) on seizure days compared to seizure-free days.** *Indicates significant difference between seizure-free and seizure distributions.

| **Participant** | **Seizure-free Bedtime (relative to mean bedtime)** | **Seizure Bedtime (relative to mean bedtime)** | **Seizure-free Waketime (relative to mean waketime)** | **Seizure Waketime (relative to mean waketime)** |
|---|---|---|---|---|
| P1 | 0.11 | -0.38 | -0.1 | 0.16 |
| P2 | 0.02 | 0.71 | -0.11 | 0.06 |
| P3 | 0.06 | -0.36 | 0.0 | -0.08 |

| | | | | |
|---|---|---|---|---|
| P4 | -0.39 | -0.19 | 0.24 | 0.14 |
| P5 | 0.02 | 0.0 | 0.02 | -0.13 |
| P6 | 0.03 | -0.18 | -0.06 | 0.32 |
| P7 | -0.02 | **0.39*** | -0.08 | **0.44*** |
| P8 | 0.01 | -0.08 | -0.11 | **0.43*** |
| P9 | 0.0 | 0.1 | -0.02 | **0.58*** |
| P10 | -0.1 | **0.23*** | 0.02 | -0.08 |
| P11 | 0.03 | 0.02 | -0.04 | 0.05 |
| P12 | -0.01 | 0.63 | -0.03 | 0.09 |
| P13 | 0.11 | -0.2 | -0.03 | 0.08 |
| P14 | 0.05 | 0.0 | -0.04 | -0.24 |
| P15 | 0.02 | **-0.33*** | 0.0 | **-0.16*** |
| P16 | 0.03 | -0.1 | 0.0 | -0.05 |
| P17 | 0.05 | -0.05 | -0.03 | 0.12 |
| P18 | 0.17 | **-1.09*** | 0.01 | -0.16 |
| P19 | -0.17 | -0.03 | 0.12 | -0.15 |
| P20 | -0.34 | **3.78*** | -0.13 | **3.5*** |
| P21 | 0.02 | -0.11 | -0.02 | -0.23 |
| P22 | 0.0 | 0.05 | 0.0 | 0.05 |
| P23 | -0.01 | **0.71*** | -0.07 | **1.3*** |
| P24 | 0.06 | -0.61 | -0.05 | **0.31*** |
| P25 | -0.02 | 0.24 | 0.07 | -0.41 |
| P26 | -0.03 | 0.57 | 0.06 | -0.33 |
| P27 | 0.07 | -0.09 | -0.06 | 0.08 |
| P28 | 0.06 | **-0.61*** | 0.04 | **-0.89*** |
| P29 | 0.1 | -0.18 | 0.21 | **-0.27*** |

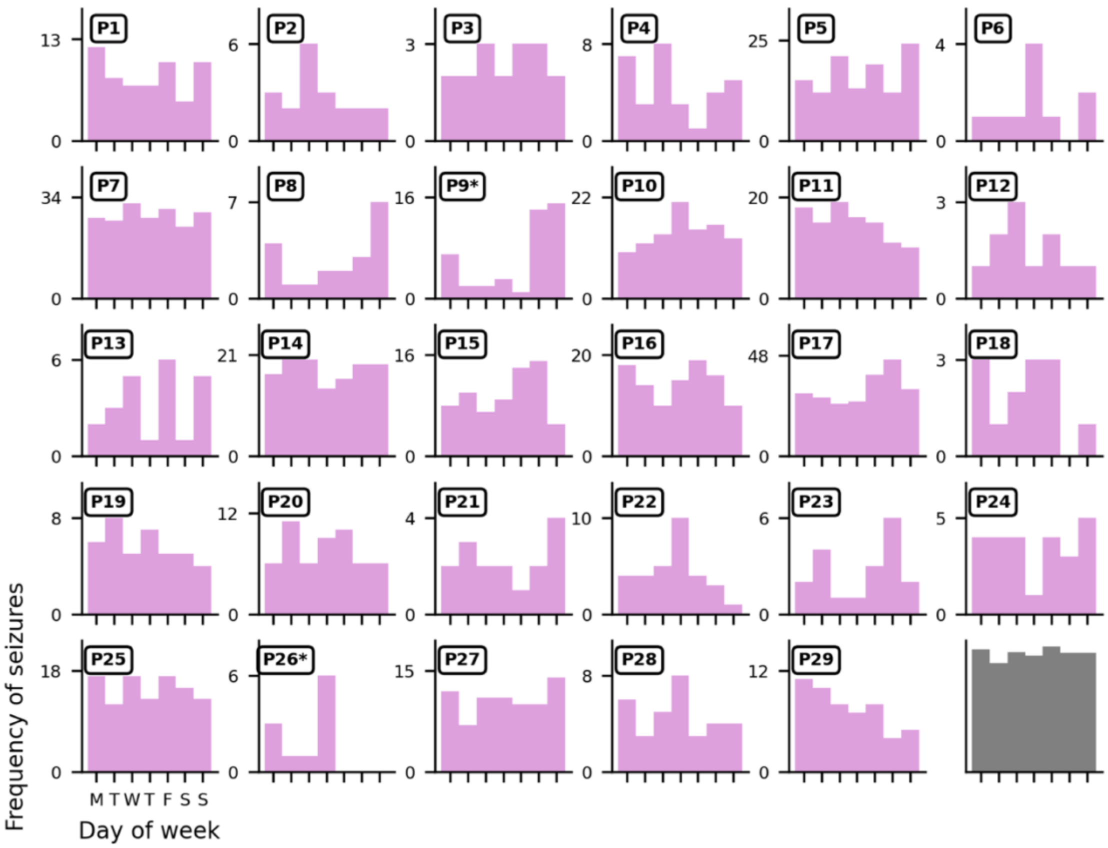


**Supplementary Figure 3. Frequency of seizure occurrence on days of the week for each participant.** * Indicates the distribution is significantly non-uniform based on the chi-squared test. Only two participants (P9 and P26) had a significantly non-uniform distribution. The grey distribution in the bottom right corner is the average distribution across all individuals.

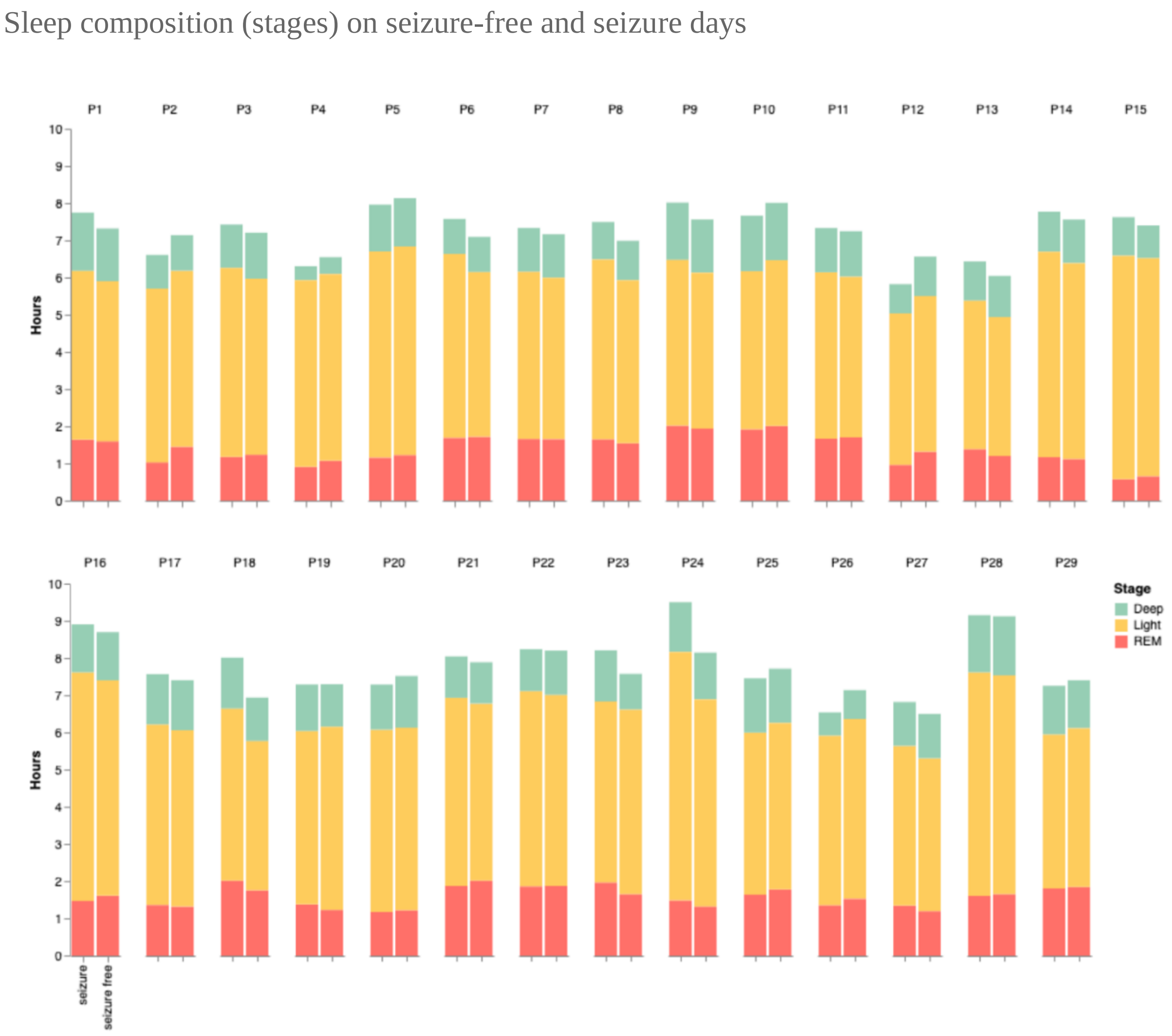


**Supplementary Figure 4. Average sleep stage (Deep, Light, REM indicated with green, yellow and red colours) breakdown during sleep prior to seizure and seizure-free days for each participant.** Participant numbers are shown above each bar graph set. Average sleep compositions for seizure days are given as the left bar graph and average sleep compositions for seizure-free days are given as the right bar graph.

**Supplementary Table 5. P-values table of sleep stage total duration and proportion the night prior to seizure days compared to seizure-free days.** P-values calculated using Wilcoxon signed rank sum test. *indicates significant association between stage and seizures.

| Participant | REM time | REM proportion | Light time | Light proportion | Deep time | Deep proportion |
|---|---|---|---|---|---|---|
| P1 | 0.425 | 0.3297 | 0.1693 | 0.9236 | **0.0269*** | 0.5919 |
| P2 | **0.015*** | **0.0139*** | 0.4745 | 0.1561 | 0.9234 | 0.7252 |

| | | | | | | |
|---|---|---|---|---|---|---|
| P3 | 0.6601 | 0.4247 | 0.0885 | 0.2455 | 0.6945 | 0.3498 |
| P4 | 0.2925 | 0.1109 | 0.5941 | 0.2164 | 0.4792 | 0.6623 |
| P5 | 0.2019 | 0.2724 | 0.3653 | 0.3354 | 0.3534 | 0.7326 |
| P6 | 0.7032 | 0.533 | **0.0285*** | 0.3866 | 0.7619 | 0.7553 |
| P7 | 0.9974 | 0.3115 | 0.1731 | 0.371 | 0.7322 | 0.7758 |
| P8 | 0.4279 | 0.9926 | 0.11 | 0.6646 | 0.7852 | 0.5155 |
| P9 | 0.1155 | 0.4152 | **0.0105*** | 0.7729 | 0.1691 | 0.8611 |
| P10 | 0.245 | 0.9996 | **0.0392*** | 0.8955 | 0.4788 | 0.8782 |
| P11 | 0.5736 | 0.1961 | 0.1272 | 0.1474 | 0.3801 | 0.3358 |
| P12 | **0.0341*** | 0.0554 | 0.5286 | **0.0312*** | 0.0522 | 0.2408 |
| P13 | 0.1574 | 0.2951 | 0.2712 | 0.9027 | 0.4131 | 0.0636 |
| P14 | 0.507 | 0.7766 | 0.2346 | 0.6151 | 0.21 | 0.0888 |
| P15 | 0.1241 | 0.0606 | 0.1631 | 0.4837 | **0.0006*** | **0.0031*** |
| P16 | **0.0012*** | **0.0001*** | **0.0019*** | **0.0038*** | 0.8926 | 0.624 |
| P17 | 0.405 | 0.5424 | 0.1476 | 0.9008 | 0.9189 | 0.4198 |
| P18 | 0.0831 | 0.7164 | **0.0414*** | 0.5088 | 0.1708 | 0.5665 |
| P19 | 0.1374 | **0.0426*** | 0.4024 | **0.0057*** | 0.1942 | 0.0617 |
| P20 | 0.3691 | 0.6841 | 0.9681 | 0.0599 | **0.0087*** | **0.014*** |
| P21 | 0.3979 | 0.0736 | 0.1446 | 0.1066 | 0.8798 | 0.7893 |
| P22 | 0.5034 | 0.7271 | 0.356 | 0.3369 | 0.6514 | 0.2488 |
| P23 | **0.0078*** | 0.0689 | 0.8298 | **0.0023*** | **0.0002*** | **0.0014*** |
| P24 | 0.2311 | 0.5945 | **0.0007*** | 0.5466 | 0.2848 | 0.7392 |
| P25 | 0.0836 | 0.1326 | 0.3282 | 0.4037 | 0.6003 | 0.3468 |
| P26 | 0.3619 | 0.8206 | 0.4847 | 0.601 | 0.2788 | 0.4423 |
| P27 | 0.0791 | 0.2 | 0.2713 | 0.7873 | 0.6285 | 0.2067 |
| P28 | 0.2699 | 0.1713 | 0.8441 | 0.2116 | 0.6945 | 0.4742 |
| P29 | 0.4407 | 0.6296 | 0.4841 | 0.9388 | 0.9665 | 0.6637 |
